# Sedentary Time Elimination with Periodic activity Snacks (STEPS): protocol for a co-designed feasibility trial in people with severe mental illness

**DOI:** 10.1101/2025.07.08.25331057

**Authors:** M Trott, U Arnautovska, N Korman, G Ritchie, D Siskind

## Abstract

**Background:** People with severe mental illness (SMI) experience substantially reduced life expectancy, largely due to cardiovascular disease (CVD). Sedentary behaviour is a major risk factor for CVD, and individuals with SMI spend significantly more time sedentary and are less physically active than the general population. While interventions to increase physical activity have focused on structured exercise, these are not accessible or acceptable to many consumers. The STEPS study aims to test and evaluate the feasibility of a co-designed novel intervention using “activity snacks” to interrupt sedentary behaviour in people with SMI.

**Methods:** The STEPS study is a single-arm feasibility trial. We will recruit 20 participants with SMI to complete a 6-week intervention. The intervention will include the use of the STEPS smartphone app to prompt short bouts of movement across the day, the type of which are personalised to the individual. Feasibility will be assessed through recruitment and retention rates, adherence to the intervention, and acceptability measured via qualitative interviews. Secondary outcomes include changes in sedentary time, physical activity, mood, and quality of life.

**Discussion:** This study will assess the feasibility and acceptability of a co-designed activity snack intervention to reduce sedentary time in people with SMI. Findings will inform the design of a future powered trial to test efficacy.

**Trial registration:** This trial is prospectively registered with the Australia New Zealand Clinical Trials Registry (ACTRN12625000265471p).

## Background and Rationale

Severe mental illnesses (SMI), including schizophrenia, bipolar affective disorder, and major depression, impact up to 4.3% of the global population(1). Compared to the general population, people with SMI have a much higher prevalence of physical comorbidities, including metabolic, respiratory, and cardiovascular disorders (2). Indeed, life expectancy in people with SMI is 15-20 years shorter than the general population (3), with cardiovascular disease (CVD) being a leading cause of death in this cohort (4, 5). In turn, one of the leading risk factors of CVD is sedentary behaviour, especially in people with SMI (6). Alarmingly, reviews reporting that people with SMI are sedentary for 12.6hrs/day, and compared to the general population, are 50% less likely to partake in 150 or more minutes of physical activity per week (7). The evidence base suggests it is, therefore, highly likely that high levels of sedentary behaviour contribute significantly to the mortality gap experienced in people with SMI and poor mental health.

Systematic reviews examining interventions aimed to reduce sedentary behaviour and increase physical activity in people with SMI found that almost all interventions have exclusively involved consumers partaking in structured exercise sessions, typically gym-based exercise prescriptions (8, 9) which presents several problems, including (a) structured exercise interventions exclude people who may not prefer this type of physical activity; (b) structured exercise programs typically rely on trained staff to deliver them; and (c) physical activity and sedentary behaviours are not mutually exclusive. Further, the association between sedentary behaviour and CVD is independent of total physical activity levels (10) and there is evidence that sedentary behaviour can also affect mood independent of physical activity levels (11), highlighting the need to consider sedentary behaviour and physical activity as separate therapeutic targets when developing novel interventions.

Recent exercise interventions have shown that recruitment for exercise-related interventions can be difficult, with over 35% of people declining because they do not want to engage in structured exercise(12). This method of intervention, therefore, *actively excludes a significant proportion of people with SMI*, is challenging to up-scale, and likely produces biased results. Furthermore, structured exercise may not be suitable to people with SMI who may not have a preference for this type of activity (which is important for internal, autonomous motivation) (13, 14), and who can experience significant motivational and cognitive deficits. Also, physical activity interventions supervised by qualified staff are a high-cost resource that may not be accessible in all settings. Given the persistence of motivational deficits in people with SMI (15), diverse interventions that may impact autonomous motivation for physical activity must be explored.

Interventions that aim to increase physical activity levels have yielded promising results, however these results do not typically extend to reducing sedentary behaviour, which is a risk factor independent of physical activity levels. For example, someone could be doing above 150mins/wk of physical activity yet exhibit long bouts of sedentary behaviour in between.

A potential solution to these problems could be in the form of physical activity ‘snacks’ – defined as bouts of physical activity under 10 minutes in duration. Whereas traditional physical activity interventions focus on discrete bouts of activity for a set time/week, physical activity snacks can be conducted at any time, in any place, without specialised clothing or equipment, and can be easily personalised to a person’s preferences. This approach requires a relatively small time commitment, and little planning. It also presents a ‘buy one get one free’ approach to physical activity – simultaneously decreasing total sedentary behaviour while increasing total physical activity, increasing its effectiveness against CVD. Indeed, research has shown interventions aiming to simultaneously decrease sedentary behaviour and increase physical activity have significant positive impacts for cardiovascular health compared to only physical activity interventions (16). Physical activity snacks have several benefits in the general population, including improved cardio-metabolic outcomes, reduced stress, depressive symptoms, increased positive mood state and decreased negative mood state (13, 17, 18). Although the feasibility and acceptability of digital interventions in people with SMI (including wrist worn devices) have been established (19), there are no interventional studies examining the feasibility and acceptability (and effectiveness) of physical activity snacks in people with SMI, despite the extremely large advantages of reducing sedentary behaviour and increasing physical activity engagement to both physical and mental health. The aim of this study, therefore, is to co-produce and test a digital app that aims to reduce sedentary behaviour through the promotion of physical activity snacks in people with SMI. Specifically, we will co-design a digital app interface and messaging service that will push messages on to a wrist-worn activity tracker at various times during the day, prompting users to partake in activity snacks.

## Methods

### Setting and Participants

Participants will be recruited from community mental health services within Metro South Health, Queensland, Australia.

### Inclusion and exclusion criteria

Inclusion criteria will include anyone with a diagnosis of either schizophrenia/schizoaffective disorder, major depression, or bi-polar disorder, who are >18years and have capacity to consent as determined by their treating clinician. They are also required to own either an Android or Apple smartphone for app downloading. Informed consent will be sought from a member of the study team, with participants able to withdraw at any time.

### Intervention

Participants will use the app and wear a wrist-worn Garmin activity tracker for six weeks. The app is designed to prompt users via the Garmin to partake in at least three (defined by the user) physical activity snacks during the day, the timing of which will be customisable to the individual. The mode of these physical activity snacks will be personalised via users choosing their preferred type of physical activity from either a pre-determined list, or other types of physical activity of their choosing. We anticipate the pre-determined list to be as follows:

– Walking up and down stairs
– Walking outside
– Housework/household chores
– Other (users will be asked to specify)

### Primary Outcomes

Feasibility will be determined by the proportion of users who consent to participate, and proportion of users who complete the study. Usability will be assessed using the System Usability Scale (SUS) (20), a 10 item questionnaire with a 5-point response scale that has shown good validity and reliability in this population (21). The scale has been found to effectively differentiate between usable and unusable systems. Motivational constructs mediating physical activity and behaviour change will be assessed using the Behavioural Regulation in Exercise Questionnaire (BREQ-3) (22).

### Secondary outcomes

Sedentary behaviour and physical activity levels will be measured by two measures: (a) the simple physical activity questionnaire (SIMPAQ), a validated physical activity measure for people with SMI (23); and (b) accelerometry metrics from the Garmin watch, which have shown comparatively accurate estimates for sedentary behaviour and physical when compared to traditional accelerometry (e.g. ActiGraph, etc) (24). The K-10 questionnaire (25) will be administered, a validated tool that measures psychological distress across all three types of SMI (26). Finally, quality of life will be assessed using the Recovering Quality of Life (ReQoL) tool, a patient-reported quality of life measure specifically developed for people with SMI (27). Other measures will include waist/hip ratio, weight, and body mass index. The Garmin device will also gather heart rate data. Adverse events will be collected at weekly phone call check ins with participants. Participants will be reimbursed with a $50 voucher at baseline, and a $50 voucher at 6 week follow up. Assessment timepoints are summarised in Figure 1.

**Figure 1.**
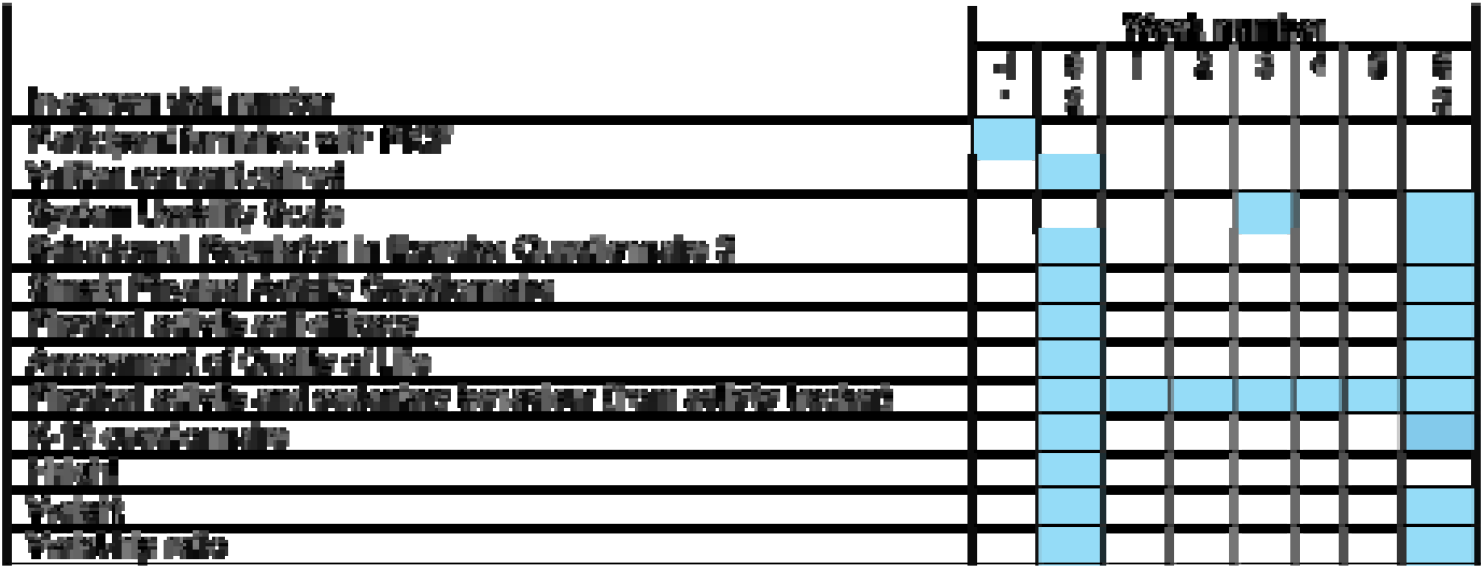
STEPS schedule.

### Sample Size

We will aim for a total sample of 20 participants, which has been informed by recommendations for usability testing of eHealth tools, (28) accounting for 10% drop out as per previous studies of digital interventions in SMI (19). Due to the single arm nature of the trial and primary outcome of feasibility and acceptability, a power calculation was not conducted. We will purposefully sample for diversity of backgrounds (in terms of SMIs, age, gender, physical health concerns), to maximise variability of experiences and needs (28). Although it is likely that this sample size will not provide statically significant results regarding the secondary outcomes, we will use the effect sizes as preliminary evidence to warrant a fully powered trial in the future.

### Statistical Analysis

All data will be managed and stored as per the National Health and Medical Research Council’s Australian Code for the Responsible Conduct of Research guidelines. The acceptability and feasibility of the intervention will be reported using descriptive statistics as means and standard deviations or number and percentages as appropriate. The qualitative data gathered from the end of intervention semi-structured interviews will be analysed using thematic analysis to identify meaningful patterns within the data based on grounded theory, facilitating a deeper and richer understanding of the participants’ experiences. All secondary (pre-post) outcomes will be analysed using either a t-test or Mann’s Whitney U test, dependent on the normality of the data. Effect sizes for secondary outcomes will be calculated as Cohen’s *d*, based on standard calculations based on mean changes and pooled standard deviations. We will also determine associations between the SIMPAQ and the Garmin physical activity and sedentary behaviour data using the Pearson’s coefficient.

### Role of the trial sponsor and funder

The trial sponsor and funders have no role in the design, collection, management, analysis, or interpretation of data. They will also have no role in writing or submitting final reports for publications.

### Dissemination

Findings will be shared with our consumer advisory group, and through peer-reviewed journal publications and academic conferences. The full protocol as approved by ethics will be publicly available via the Australia New Zealand Clinical Trials Registry. Individual patient data will be available from the corresponding author upon reasonable request.

### Discussion

The STEPS study will assess the feasibility and acceptability of a co-designed activity snack app with wrist watch, to reduce sedentary time in people with SMI. There is currently limited research focused on reducing sedentary behaviour in this population, despite its known links to poor physical and mental health outcomes. Findings from this study will be assessed against pre-specified feasibility and acceptability criteria to inform the design of a future fully powered trial. The intervention will target a clinically important and underserved population who spend prolonged periods sedentary and face significant barriers to engaging in structured exercise. If feasible and acceptable, the STEPS intervention could offer an accessible and low-burden alternative to conventional physical activity programs, with the potential to reduce cardiometabolic risk and improve quality of life. Additional benefits may include improved mental health, greater daily structure and engagement, and the potential for digital delivery to enhance scalability across community mental health services.

Moreover, the integration of user-centred co-design and digital prompts tailored to individual preferences may enhance user engagement and sustainability. Understanding the factors that influence uptake and adherence — such as usability, motivation, and support — will provide valuable insights for optimising the intervention and informing implementation strategies.

### Trial Status

This protocol is version 1.0, dated 5 March 2025. Ethics approval is approved by the Metro South Human Research Ethics Committee (HREC/2025/QMS/117193). Recruitment is expected to commence in January 2026 and will continue for approximately six months. This trial is registered with the Australia New Zealand Clinical Trials Registry (ACTRN12625000265471p). Trial is sponsored by Metro South Addiction and Mental Health Services. Any protocol changes/deviations will be reported to the appropriate ethics committee, and to the Australia New Zealand Clinical Trials Registry.

## Data Availability

All data produced in the present study are available upon reasonable request to the authors

## Abbreviations

ANZCTR: Australian New Zealand Clinical Trials Registry
BREQ-3: Behavioural Regulation in Exercise Questionnaire – Version 3
CVD: Cardiovascular Disease
K-10: Kessler Psychological Distress Scale
NHMRC: National Health and Medical Research Council
ReQoL: Recovering Quality of Life
SIMPAQ: Simple Physical Activity Questionnaire
SMI: Severe Mental Illness
STEPS: Sedentary Time Elimination with Periodic activity Snacks
SUS: System Usability Scale

## Declarations

### Ethics approval and consent to participate

Ethics approval is approved by the Metro South Human Research Ethics Committee (HREC/2025/QMS/117193). Full informed consent will be sought for every participant.

### Consent for publication

Not applicable

### Availability of data and materials

Not applicable

### Authors’ contributions

MT conceived the study, gained funding, and wrote the protocol. UA, NK, GR all contributed to the writing of the manuscript. DS contributed to the conceptualisation and writing of the manuscript in a senior, supervisory role.

## Acknowledgements

Not applicable

